# Disease Outcomes in Boys with *ABCD1* Variants Identified by Newborn Screening for X-ALD

**DOI:** 10.64898/2026.06.30.26356979

**Authors:** Cecilie S. Videbaek, Danielle HJ Kim, Hannah S. Hart, Robert Thompson, Razina Aziz-Bose, Lachelle Purnell-Savoy, Sonum Bharill, Ezzat Hashemi, Joseph Orsini, Elisa Seeger, Miranda McAuliffe, Isha Srivastava, Jennifer A. MacLean, Sejal Shah, Ali Fatemi, Julie S. Cohen, Eric Mallack, Troy Lund, Florian Eichler, Joshua L. Bonkowsky, Laura Adang, Zihuai He, Allan M. Lund, Keith P. van Haren

## Abstract

**Objectives:** To determine whether boys with VUS detected through Newborn screening (NBS) for Adrenoleukodystrophy (ALD) develop adrenal insufficiency (aiALD) and cerebral ALD (cALD) at rates comparable to those with pathogenic variants, and to evaluate the relationship between C26:0-lysophosphatidylcholine (C26:0-LPC) levels and clinical outcomes.

**Methods:** We conducted a retrospective multicenter cohort study (2013–2025) across six US centers, including 201 males identified through NBS in 19 states. Variants were classified as pathogenic (n=65), likely pathogenic (n=45), or VUS (n=88). Primary outcomes were development of aiALD and cALD; secondary outcomes included C26:0-LPC levels. Statistical analyses included Kaplan–Meier, mixed-effects regression, and Cox models.

**Results:** 201 males with *ABCD1* variants identified through NBS for ALD. Median age at last follow-up was 4.2 years (IQR 2.5–7.9). Overall, 26% developed aiALD (54% pathogenic, 16% likely pathogenic, 11% VUS), and 8% developed cALD (11%, 9%, and 4.5%, respectively). Pathogenic/likely pathogenic variants were associated with higher odds of aiALD than VUS (OR 5.8; 95% CI 2.16–15.58; p=0.001). At 150 months, 39% of individuals with pathogenic/likely pathogenic variants remained free of aiALD versus 85% with VUS. C26:0-LPC levels were higher in pathogenic variants and correlated with genotype (p=0.0006). Higher levels were associated with increased aiALD risk and earlier onset (HR 1.38 per 0.1 µmol/L; 95% CI 1.20–1.59; p<0.0001).

**Conclusions:** Boys with VUS had lower rates of aiALD and lower C26:0-LPC levels than those with pathogenic variants, although some developed disease. C26:0-LPC correlates with genotype and risk, supporting its role in variant classification and risk-stratified surveillance.

**What’s Known on This Subject:** Newborn screening for X-linked adrenoleukodystrophy has increased identification of variants of uncertain significance, accounting for up to 50% of screen-positive cases in some states. These are often associated with borderline biomarker levels, and their natural history remains poorly understood.

**What This Study Adds:** Screen-positive individuals with VUS had substantially lower rates of disease onset than those with pathogenic variants. Newborn biomarker levels also correlated with variant pathogenicity and disease onset, which may aid future variant classification and risk stratification.

## Background

In 2024, nearly all 3.6 million infants born in the US were screened for X-linked adrenoleukodystrophy (ALD), a rare metabolic disorder with potentially deadly endocrine and neurologic manifestations that are treatable if identified early.^1^ ALD is caused by pathogenic variants in the *ABCD1* gene, leading to the accumulation of very-long-chain fatty acids (VLCFAs) in adrenal glands, brain myelin, and other tissues^2^.

While natural history studies have reported incidences of ALD ranging from 0.5 to 6 per 100,000 males,^3–9^ newborn screening (NBS) programs have suggested much higher incidences^10^. Phenotypic penetrance varies by age and sex, with earlier symptom onset in males. Historical cohorts suggest that more than half of males with pathogenic *ABCD1* variants will develop adrenal insufficiency (aiALD) before adulthood, and about one-third of males will develop inflammatory cerebral demyelination (cALD) by the age of 10^1,2^. Adrenal insufficiency is readily treated with adrenal replacement therapy, while cALD requires riskier treatments with hematopoietic stem cell transplantation (HSCT), including Skysona, an FDA-approved *ex vivo* gene therapy^11^. In adulthood, most individuals with pathogenic *ABCD1* variants develop progressive adrenomyeloneuropathy (AMN), for which no therapies currently exist.^12,13,14^ Currently, there is no established genotype-phenotype correlation in ALD, and predicting the disease course remains uncertain. Several biomarkers are being investigated as predictors of ALD phenotype onset and progression, among which C26:0-lysophosphatidylcholine (C26:0-LPC) remains the most promising.^15,16,17,18^

Both aiALD and cALD are life-threatening without treatment, and the success of treatment relies on early intervention.^1,19^ Therefore, presymptomatic surveillance is crucial in the management of ALD and involves serial laboratory assessment of adrenal function for aiALD every 3-12 months and regular brain MRI studies for cALD every 6-12 months.^20^

Individuals at risk for ALD symptoms can be diagnosed presymptomatically through cascade testing originating from a symptomatic family member or a positive NBS result. NBS for ALD is implemented in 47 American states, the Netherlands^21^ and Taiwan^22^, and uses mass spectrometry to measure C26:0-LPC in dried bloodspots,^23–25^ followed by sequencing of the *ABCD1* gene.^10^ Genetic variants are classified into one of five ACMG/AMP categories based on known or predicted pathogenicity: benign, likely benign, variant of uncertain significance (VUS), likely pathogenic (LP), or pathogenic (P). Although most clinical laboratories apply ACMG/AMP interpretation guidelines, laboratory-specific adaptations or evidence weighting can produce discordant classifications.^26–28^ Follow-up of 32 boys identified via ALD NBS in Minnesota and followed for a median of 5 years suggests NBS may identify boys with lower disease risk than historical cohorts with symptomatic diagnosis of ALD.^29^

Since 2013, when New York state became the first government to implement universal NBS for ALD,^30^ the growing adoption of ALD NBS has led to increased identification of individuals with *ABCD1* VUS.^31,32^ Individuals with these variants often exhibit modest elevations in C26:0-LPC and no family history of ALD, complicating traditional diagnostic and prognostic guidance.^20^ NBS for ALD is unique in that a positive result leads to lifelong surveillance rather than immediate treatment.^20,33,34^ While early identification enables intervention before irreversible disease, it places a long-term psychological and logistical burden on families^35^ and healthcare systems.^36^ Therefore, accurate identification of individuals truly at risk for symptomatic ALD is crucial.^37^ Efforts are underway to improve the prediction of *ABCD1* variant pathogenicity, but all require high-quality natural history data.^38,39^

We sought to determine whether boys with VUS identified via NBS developed ALD-associated symptoms at comparable rates to those with pathogenic or LP (P/LP) variants. To investigate this, we conducted a multi-site retrospective cohort study comparing rates of aiALD and cALD between boys with VUS versus P/LP *ABCD1* variants, and examined relationships between symptoms, variant classification, genotype, and C26:0-LPC levels.

## Methods

### Study design

This cohort study was performed according to the STROBE statement.^40^ Retrospective data from 2013-2025 were collected from October 2024 to July 2025 across six US medical centers: Stanford University (SU), Children’s Hospital of Philadelphia (CHOP), Massachusetts General Hospital (MGH), University of Utah (UU)/Primary Children’s Hospital, University of Minnesota (UM), and the Kennedy Krieger Institute (KKI). Data was collected via chart review.

### Protocol approvals

The work and data collection for this study was covered by IRB protocols (SU #59014; CHOP #20-018050 and 14-011236; MGH #2012P000132; UU #19596; KKI #496013). Approvals for data collection at UM are described elsewhere^29^. The study complied with HIPAA regulations for data protection.

### NBS methods across states

All individuals were identified presymptomatically via NBS positive screens for ALD across 19 US states. All states utilize C26:0-LPC levels and *ABCD1* sequencing for baseline screening, though methodologies and thresholds vary by state (**Supplemental Table 1**).

### Study population

#### Inclusion criteria

Inclusion criteria were (1) male sex with (2) an *ABCD1* genetic variant classified as either VUS, LP, or pathogenic that was (3) associated with an elevated C26:0-LPC level and (4) identified either directly via NBS or indirectly via cascade testing that originated from a family member identified through NBS. For individuals identified via cascade testing, we required biochemical confirmation of a C26:0 level above the normal range specified by the testing laboratory.

#### Identification of study subjects

Subjects were identified via chart review using diagnostic codes, NBS referral records, and center-specific ALD registries^29^.

### Study variables and definitions of aiALD and cALD

Data variables included *ABCD1* genotype, variant classification at screening, C26:0-LPC levels from NBS in µmol/L, reason for diagnosis (NBS versus cascade testing prompted by NBS). Clinical or biochemical evidence of adrenal insufficiency was required for aiALD^34^. We applied an inclusive definition of aiALD that encompassed both full adrenal insufficiency, which requires daily corticosteroid use, as well as partial adrenal insufficiency, which typically prescribes corticosteroid use only during periods of physiological stress. Age at onset of aiALD was defined as date first prescribed steroids for either partial or complete adrenal insufficiency. Age at onset of cALD was defined as MRI with a Loes score^41^ ≥0.5 with gadolinium enhancement or subsequent progression. Age at last follow-up was the date of the last clinical encounter.

For analysis of *ABCD1* variant classification, we used the classification reported by the sequencing laboratory at the time of diagnosis. Separately, we recorded the current variant classification (as of May 2026) from ClinVar^42^ and the *ABCD*1-registry^31^.

For biochemical markers, we used C26:0-LPC levels measured by LC-MS on dried newborn blood spots from the child’s first heel prick. For most states, this is the second tier of the NBS (**Supplemental Table 1**).

### Statistics

In our analyses, LP/P variants were grouped together as they are similar within the ACMG classification system compared to VUS. To compare group differences in C26:0-LPC levels, we tested for normality and used either Mann-Whitney (non-normal distributions) or Welch’s t-test (normal) where appropriate.

Kaplan-Meier curves were used to estimate survival probabilities for aiALD and cALD by variant classification. Time-to-event was defined as age at onset, with follow-up from birth to event or censoring at 150 months (RStudio, prodlim package).

We converted all C26:0-LPC values to µmol/L and scaled per 0.1 µmol/L increase. We used a logistic mixed-effects regression to assess associations of *ABCD1* variant classification and C26:0-LPC levels with aiALD, including a random intercept for family, and reported the results as an odds ratio (OR). We used a Cox proportional hazards model with a shared frailty term for family to evaluate associations of variant classification and C26:0-LPC levels with time to aiALD onset and reported the results as a hazard ratio (HR) and 95% confidence intervals (CIs). We used a linear mixed-effects model to assess the association between *ABCD1* genotype and C26:0-LPC levels, with genotype as a fixed effect and family as a random effect. We estimated least squares means (95% CIs) using c.839G>A as reference. We used SAS OnDemand 9.4.

Robust modeling for cALD-C26:0-LPC association was not feasible due to the limited number of individuals who developed cALD.

We used Type III tests to evaluate overall effects; p<0.05 was considered statistically significant, and missing data were coded as not available.

## Results

Among 379 potential subjects identified across all centers, 201 individuals met inclusion criteria, which included NBS positives screens from 19 different US states. **Table 1** provides an overview of our cohorts and outcomes. Median age at last follow-up was 4.2 years (IQR 2.5-7.9), 44% of our cohort were individuals with a VUS. By the time of last follow-up, 52 individuals (26%) developed aiALD and 16 (8%) had developed cALD.

**Table 1:** Schematic overview of demographic and outcome data. Patients were placed in either P, LP, or VUS group based on the classification of their genetic variant as it was reported by the newborn screening center. Original variant classifications were not available for three patients (**), two with aiALD (***) and one with cALD (****). IQR= Interquartile range

|  | Total | Pathogenic | Likely pathogenic | VUS |
| --- | --- | --- | --- | --- |
| **n (% of total cohort)** |  |  |  |  |
| Stanford | 60 (30%) | 20 | 11 | 29 |
| Children's Hospital of Philadelphia | 45 (22%) | 15 | 6 | 24 |
| Minnesota | 31 (15%) | 8 | 16 | 7 |
| Utah | 32 (16%) | 11 | 5 | 16 |
| Massachusetts General Hospital | 20 (10%)** | 5 | 4 | 8 |
| Kennedy Krieger Institute | 13 (6%) | 6 | 3 | 4 |
| Total | 201 | 65 (32%) | 45 (22%) | 88 (44%) |
| **Median age (IQR) at last follow-up (years)** |  |  |  |  |
| Stanford | 4.1 (2.38-7.02) | 4.5 (2.83-6.83) | 2.25 (1.37-7.58) | 4.25 (3.08-7.58) |
| Children's Hospital of Philadelphia | 3.58 (0.75-5.75) | 1.41 (0.5-4.79) | 1.08 (0.46-4.02) | 4.13 (1.3-6.10) |
| Minnesota | 4 (3-5.5) | 3.0 (3.0-5.0) | 5 (4.0-6.25) | 3 (2.5-4.0) |
| Utah | 3.5 (1.06-17.4) | 2 (0.96-17.5) | 8.0 (3.33 – 10.25) | 3.83 (1.40-17.38) |
| Massachusetts General Hospital | 5.38 (3.65-7.06) | 4.50 (3.67-10.50) | 3.21 (1.06-6.19) | 5.79 (5.21-11.35) |
| Kennedy Krieger Institute | 4 (1.42-5.08) | 3.58 (1.85-4.75) | 5.08 (4.46-6.04) | 3.12 (1.10-6.02) |
| Total | 4.2 (2.5-7.92) | 3.96 (2.1-6.81) | 4 (2.0-7.0) | 4.21 (2.44-6.81) |
| **Median C26:0-LPC levels (IQR) from NBS (μmol/L)** |  |  |  |  |
| Total | 0.35 (0.27-0.64)<br>*57 missing | 0.80 (0.594-0.962)<br>* 25 missing | 0.33 (0.265-0.52)<br>* 10 missing | 0.29 (0.242-0.363)<br>* 20 missing |
| **n Adrenal insufficiency** |  |  |  |  |
| Total | 52 of 201 (26%)* | 35 of 65 (54%) | 7 of 45 (16%) | 8 of 88 (9%) |
| **n Cerebral ALD** |  |  |  |  |
| Total | 16 of 201 (8%)* | 7 of 65 (11%) | 4 of 45 (9%) | 4 of 88 (4.5%) |

### Variant Classification and Phenotype

We identified 99 distinct *ABCD1* variants in our cohort. **Supplemental Table 2** lists designated classifications at time of diagnosis, as well as updated classifications in the *ABCD1* Registry^31^ and ClinVar^42^ as of May 2026. Ten variants received conflicting classifications across time and state of screening. Thirty-four genotypes showed discrepancies across databases. In total, 11% (n=10) of individuals with a genotype originally classified as a VUS developed aiALD (n=8) and/or cALD (n=4) during the observation period. These clinical outcomes indicate that these 10 variants can be disease-causing and reclassification to LP status could be considered (**Table 2**).

**Table 2:** Ten individuals with ABCD1 variants that were originally classified as VUS and developed aiALD and/or cALD during the study period. Newborn C26:0-LPC levels are not available for the two individuals who were diagnosed via cascade screening. Each row represents a single case with phenotype at last follow-up.

| Variant classification in report | Variant classification in ABCD1 registry | Coding change | Amino acid change | C26:0-LPC | aiALD only | cALD only | aiALD & cALD |
| --- | --- | --- | --- | --- | --- | --- | --- |
| VUS | P | c.251C>T | p.(Pro84Leu) | 0.63 | X |  |  |
| VUS | VUS | c.452T>C | p.(Ile151Thr) | - | X |  |  |
| VUS | VUS | c.794T>C | p.(Phe265Ser) | 0.43 | X |  |  |
| VUS | LP | c.824G>C | p.(Arg275Pro) | 0.86 |  |  | X |
| VUS | LP | c.851C>T | p.(Ser284Leu) | 0.31 | X |  |  |
| VUS | VUS | c.1622A>G | p.(Tyr541Cys) | 0.185 |  | X |  |
| VUS | - | c.1652G>T | p.(Gly551Val) | 0.85 | X |  |  |
| VUS | P | c.1684T>C | p.(Ser562Pro) | 0.31 | X |  |  |
| VUS | P | c.1832A>G | p.(Gln611Arg) | 0.64 |  | X |  |
| VUS | LP | c.2000A>G | p.(His667Arg) | - |  |  | X |

Of the 52 subjects in our cohort who had developed aiALD, 35 (67%) had pathogenic variants, 7 (13%) had LP variants, and 8 (15%) had VUS (classification was unavailable for n=2). P/LP variant status was significantly associated with increased odds of aiALD compared with VUS (OR = 5.8, 95% CI: 2.16–15.58, p = 0.001). **Table 2** summarizes VUS associated with disease in our cohort. Among the aiALD patients with a VUS, only three variants (c.1184C>A (p.A395V), c.1652G>T (p.G551V), and c.794T>C (p.F265S)) were consistently classified as VUS across the online databases,^31,42^, and our dataset. One variant (c.1747G>A (p.V583M)) appeared as a VUS, LP, and P variant in our cohort (**Supplemental Table 2**).

Of the 16 subjects who developed cALD, seven (11%) had pathogenic variants, four (9%) had LP, and four (4.5%) had VUS. The limited number of cALD cases precluded robust statistical modeling of risk. Original classification was unavailable for one cALD patient. Of these four VUS, one (c.1622A>G (p.Y541C)) was consistently classified as a VUS across the *ABCD1* registry,^31^ ClinVar^42^, and our data, but none of them appeared with conflicting classifications within our cohort (**Supplemental Table 2**).

Figure 1 shows the Kaplan-Meier curve for aiALD onset in the groups with P/LP variants versus VUS. At 150 months of age, the probability of remaining free of aiALD among those with a P/LP variant was 39% (95% CI: 24%-54%) compared to 85% (95% CI: 72%-96%) in the VUS group. The probability of remaining free of cALD at 150 months was 75% (95% CI: 62%-89%) among individuals with a P/LP variant compared to 81% (95% CI: 59%-100%) in the VUS group.

**Figure 1:**
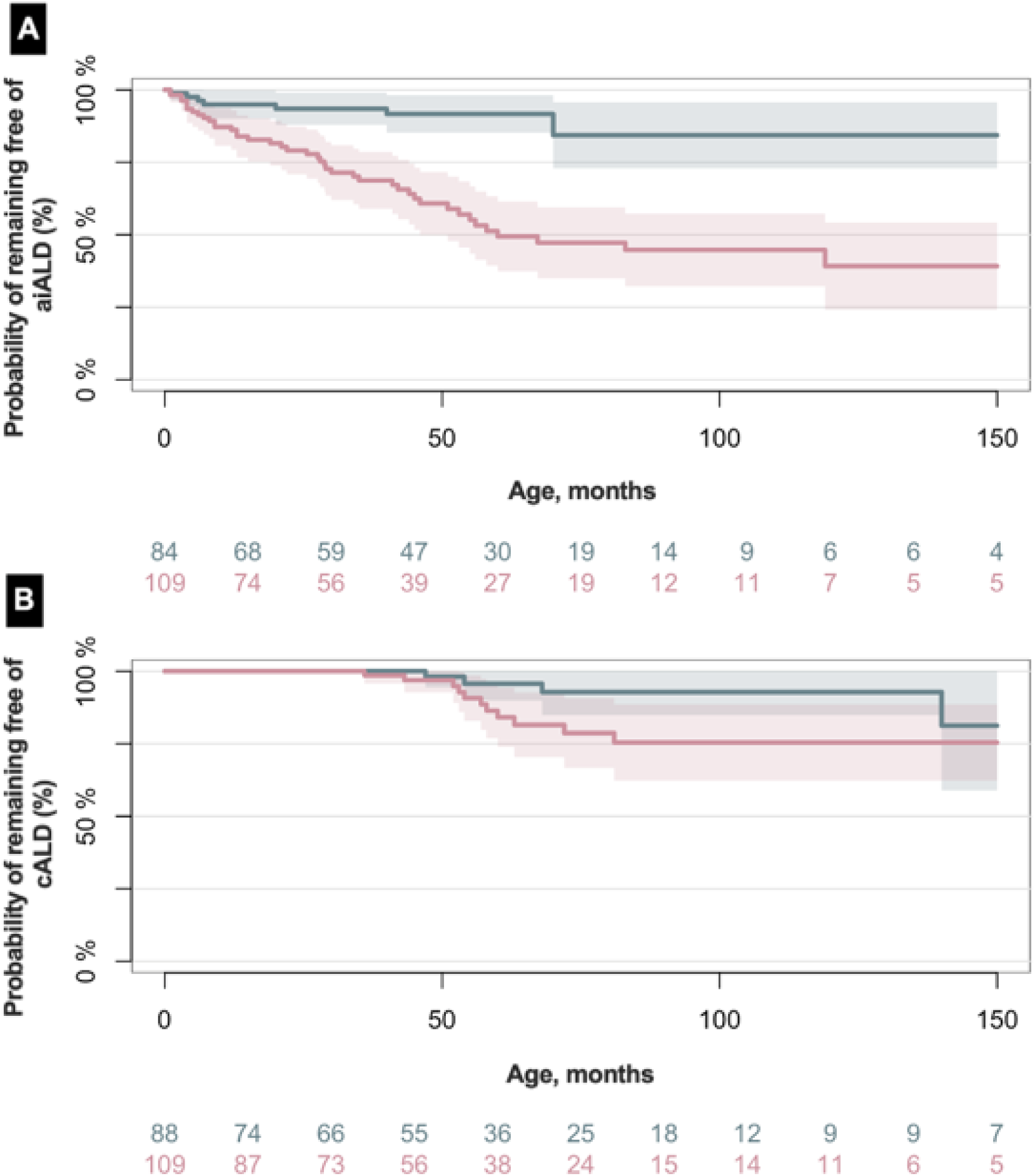
Kaplan–Meier curves illustrating that boys with ABCD1 variants classified as VUS had a higher probability of remaining free of aiALD at 150 months (82%) than those with P/LP variants (39%) (A). No significant difference in cALD risk was observed between groups (B).

### C26:0-LPC levels, Variants, and Outcomes

Newborn C26:0-LPC levels were available for 144 individuals. C26:0-LPC levels were higher among individuals with pathogenic and LP *ABCD1* variants compared with individuals with VUS where the median (IQR) was 0.80 µmol/L (0.60-1.0), 0.33 µmol/L (0.27-0.52), 0.29 µmol/L (0.25-0.37), respectively (**Table 1**; Figure 2A). Consistent with these findings, model-based estimates using adjusted means also demonstrated significantly higher C26:0-LPC levels among individuals with P/LP variants compared with VUS. Among individuals with the same variant classification, C26:0-LPC levels were higher among those who developed symptoms (Figure 2B) and were highest among those who manifested cALD (Figure 2C).

**Figure 2:**
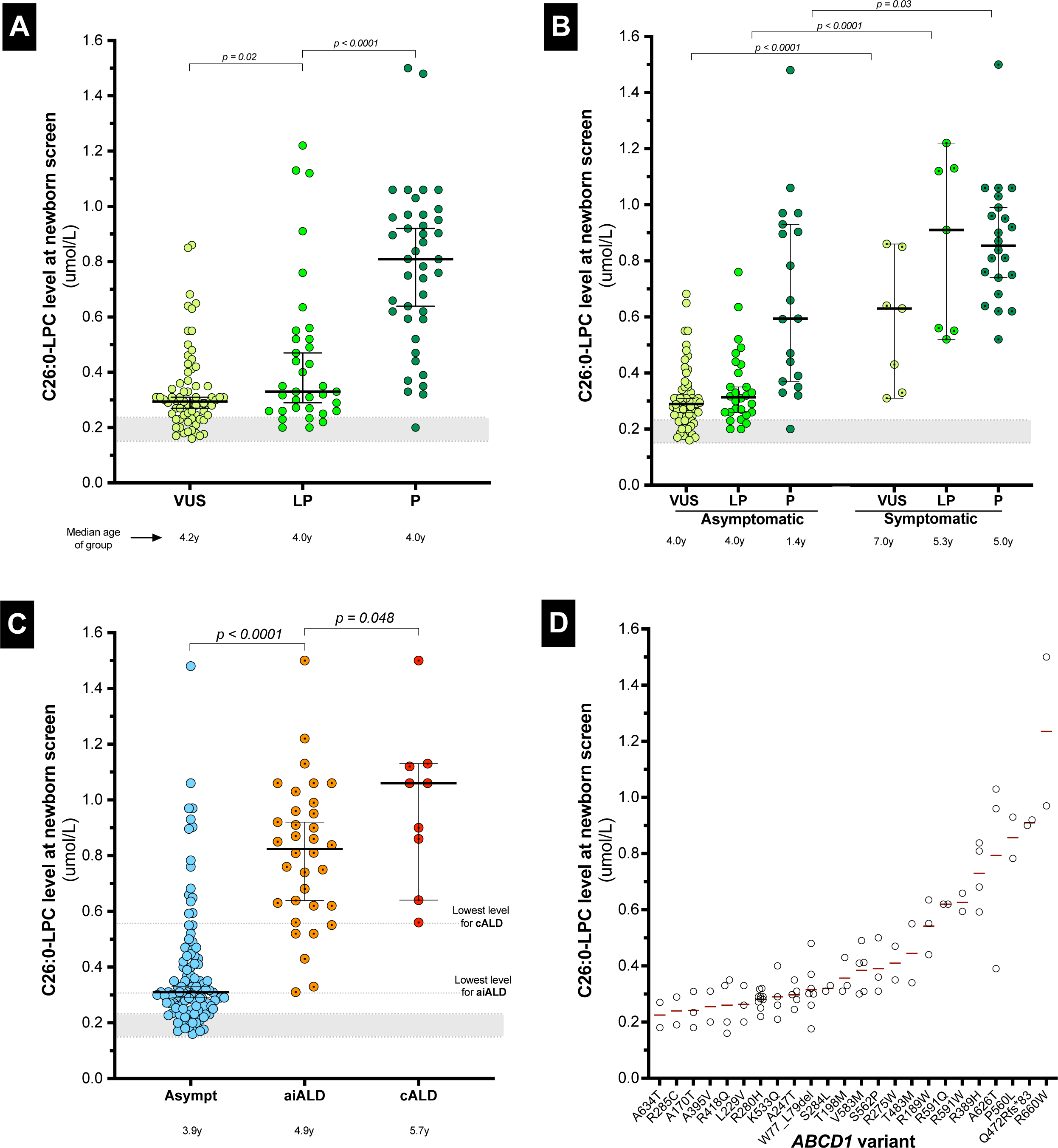
C26:0-LPC (µmol/L) at birth levels by variant classification, outcome, and genotype. Levels were higher in pathogenic/likely pathogenic variants than VUS (A) and in symptomatic versus asymptomatic individuals (B). Levels were highest in cALD (C). Similar genotypes showed consistent levels across laboratories (p=0.0006) (D). Shaded area indicates current NBS thresholds (0.15–0.25 µmol/L).

We also observed that individuals with the same genotype tended to have similar C26:0-LPC levels (Figure 2D), even across different state-run screening labs (**Supplemental** Figure 1). Our linear-mixed effects model showed that individuals with the same genotype exhibited similar C26:0-LPC levels (p=0.0006).

Among individuals who developed aiALD, the median C26:0-LPC level was 0.81 µmol/L (range: 0.31-1.5 µmol/L). Each 0.1 µmol/L increase in C26:0-LPC was associated with increased aiALD risk and earlier onset (HR = 1.38, 95% CI: 1.20–1.59, p < 0.0001).

Among individuals who developed cALD, the median C26:0-LPC was 1.06 µmol/L (range: 0.56-1.5 µmol/L); the lowest associated level was 0.56 μmol/L.

### Developmental Timing of Disease Onset

aiALD presented earlier than cALD at a median age (IQR) of 1.79 years (0.58-3.69) and 4.63 years (4.27-5.7), respectively (Figure 3A; p<0.0001). Among the 16 individuals who developed cALD, 13 (81.3%) also manifested aiALD, with aiALD preceding cALD by a median (IQR) of 3.25 years (1.24-4.21) (Figure 3B**, 3C**). Higher C26:0-LPC was associated with earlier aiALD onset, consistent with Cox model results (**Supplemental** Figure 2). Median (IQR) age among those with no symptoms at last follow-up was 3.88 years (1.35-6.75), indicating a large portion of the cohort had not yet reached the developmental window at highest risk for cALD.

**Figure 3:**
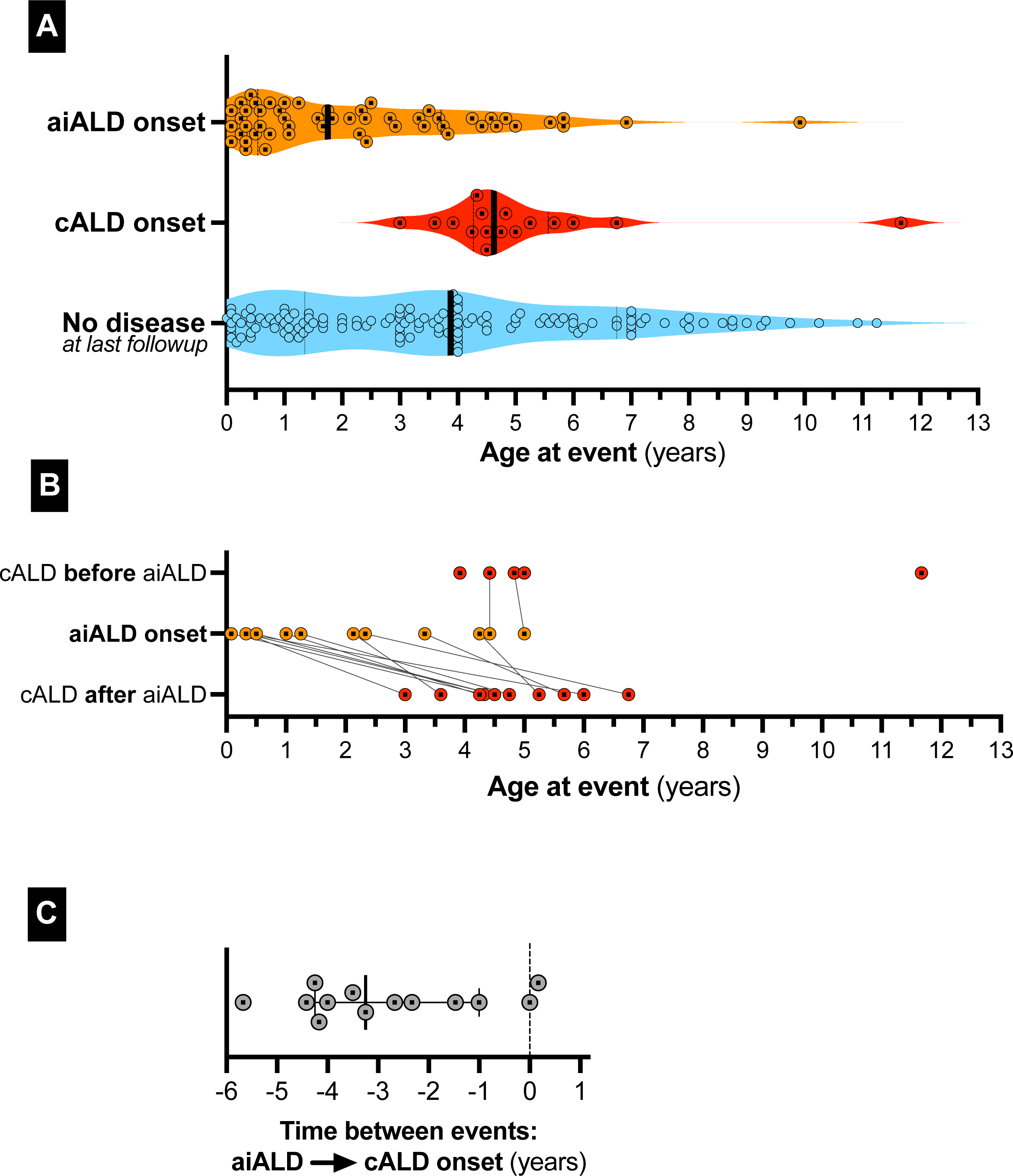
Individuals developed aiALD (orange) earlier than cALD (red), with median (IQR) ages of 1.79 (0.58–3.69) and 4.63 (4.27–5.7) years, respectively (p<0.0001). Among asymptomatic individuals, median age was 3.88 years. Most cALD cases (81%) were preceded by aiALD, with a median interval of 3.25 years.

## Discussion

In this multicenter cohort of 201 boys identified through ALD NBS, we found that variant classification and C26:0-LPC levels were both associated with the development of aiALD. We observed a substantial difference in aiALD rates between individuals with P (54%), LP (16%), and VUS (9%) variants, although our median follow-up of 4.2 years for each group limits conclusions about lifetime risk. In contrast, rates of cALD onset were lower (8% overall) and were statistically similar across genotype classes (11%, 9%, and 4.5%, respectively). We also found that newborn C26:0-LPC levels were similar among individuals with the same *ABCD1* genotype variants and were correlated with both risk and timing of aiALD onset.

Our findings indicate that individuals with a VUS are either less likely to develop aiALD compared to those with P/LP variants or that they develop symptoms later. Among individuals with P/LP variants, we observed a higher proportion of aiALD at younger ages than reported in historical natural history studies^43^. This likely reflects earlier diagnosis enabled by NBS, as seen in recent studies^29^, rather than a true increase in disease incidence. In contrast, we observed no significant differences in cALD rates between variant classification groups, likely due to the lower incidence and later onset than aiALD^3,5,7–9,44–48^. Historical cohorts suggest cALD affects 30-40% of boys by age 10 years; whether NBS-identified individuals with P variants will reach similar rates requires longer follow-up. Importantly, a subset of individuals with VUS also developed aiALD and/or cALD, underscoring the value of NBS in enabling timely intervention even when variant pathogenicity is uncertain. Although a single symptomatic case does not automatically enable reclassification, these variants can now be considered for reclassification.

Our findings support prior observations that C26:0-LPC correlates with disease manifestations in ALD and provide a quantitative estimate of risk in an NBS population^17,18^. Although C26:0-LPC levels differed by variant classification, interquartile ranges overlapped. Two individuals with aiALD had levels below 0.4 µmol/L, indicating that relatively low levels do not exclude disease risk. Among those who developed cALD, C26:0-LPC levels were higher, and disease onset occurred at older ages, typically after onset of aiALD. The association between C26:0-LPC levels and the risk of developing aiALD (hazard ratio = 1.38 per 0.1 µmol/L) offers an estimate that can inform future screening thresholds, surveillance intensity, and clinical trial design. It also supports incorporating C26:0-LPC into diagnostic panels for ALD^49^.

We observed an unexpected correlation between specific *ABCD1* genotypes and C26:0-LPC levels, consistent across states, suggesting that individual variants directly influence biochemical phenotype, possibly reflecting their functional impact on transporter efficiency. These results provide reference data for *in vitro* study of ALDP function and may prove valuable for variant classification, although further research is required before clinical application.

These findings could have direct implications for NBS programs and clinical surveillance strategies. Most states use second-tier C26:0-LPC thresholds between 0.15-0.22 μmol/L^10^. Several groups have suggested raising this threshold in order to lower the number of VUS and false positives, which could reduce the psychological, medical, and financial burden of false-positive results, but may increase the risk of missed diagnoses^18,32,50^.

As ALD NBS expands, novel *ABCD1* variants will continue to be identified, requiring careful classification. The varying phenotypes and penetrance of ALD challenge the traditional ACMG variant classification system and may require a dedicated algorithm. The ClinGen Peroxisomal Disorders Variant Curation Expert Panel (VCEP) has recently developed disease-specific criteria specifications for *ABCD1* variant classification, which will refine and standardize interpretation across laboratories^51^. Unfortunately, the most important and difficult aspect of *ABCD1* classification involves determining which *ABCD1* variants can be downgraded to a benign category or otherwise safely triaged to a less burdensome ALD surveillance strategy. Our data suggest that C26:0-LPC thresholds could potentially aid this process, but substantial work remains to understand the risks and benefits.

Several limitations should be considered. First, heterogeneous NBS protocols across states may affect C26:0-LPC comparability. Second, the median follow-up of 4.2 years is short relative to the natural history of ALD. Third, we used original variant classifications rather than updated ones; reclassification typically occurs when variants become associated with symptoms, so using current classifications would risk circular reasoning. To ensure transparency, **Supplemental Table 2** shows each genotype’s classification across sources. Fourth, our inclusive aiALD definition, encompassing both partial and complete insufficiency, may have increased the observed proportion. Finally, we included only NBS-identified individuals and excluded family members diagnosed symptomatically, so findings may not generalize to all ALD patients.

While it is still early to assess the long-term impact of ALD NBS, our findings indicate that variant classification and C26:0-LPC levels may offer valuable prognostic information that could eventually guide risk-stratified surveillance. Boys with variants originally classified as VUS had significantly lower rates of aiALD than those with LP/P variants, though some still developed disease, underscoring the need for continued vigilance. Development of aiALD was also found to correlate with C26:0-LPC levels. The correlation between specific *ABCD1* genotypes and C26:0-LPC levels raises the possibility that this biomarker provides complementary prognostic information beyond a variant’s ACMG classification, which establishes disease-causing potential but does not predict penetrance or timing of clinical manifestations in individual patients^37^. Future studies should validate these associations in larger cohorts with longer follow-up, particularly through the peak window of cALD risk. Standardizing C26:0-LPC measurement and establishing evidence-based thresholds for risk stratification could reduce the burden on families while preserving the life-saving benefits of early detection.

### Data availability statement

Individual-level data cannot be shared due to PHI restrictions. Deidentified aggregate data may be available from the corresponding author upon request and subject to appropriate approvals

## Data Availability

All data produced in the present study are available upon reasonable request to the authors

## Acknowledgements

The authors gratefully acknowledge the *ABCD1* Variant Registry and its contributors for maintaining an invaluable resource for *ABCD1* variant interpretation.

## Conflict of Interest Disclosures (includes financial disclosures): Conflicts of interest

Cecilie S. Videbæk has received travel support from Sanofi.

Danielle HJ Kim has received research funding through a Clinical Research Training Scholarship in ALD funded by Arrivederci ALD, the American Brain Foundation, and the American Academy of Neurology.

Isha Srivastava, Ali Fatemi, Florian Eichler, Joshua L. Bonkowsky, Laura Adang, and Keith P. van Haren have received research funding from the National Institutes of Health (NIH U54NS115052).

Eric J. Mallack receives research support from the National Institute of Neurological Disorders and Stroke (K23NS118044); Kennedy Krieger Institute holds institutional contracts with Minoryx Therapeutics, Ionis Therapeutics, and the N-Lorem Foundation.

Joshua L. Bonkowsky reports involvement in clinical trials with Spur Therapeutics, Calico, and Ionis; consulting for Ionis; authorship of content for UpToDate; stock ownership in Orchard Therapeutics; and receipt of royalties from BioFire.

Keith P. van Haren reports clinical trial funding from Minoryx and bluebird bio; research funding from NIH, Minoryx, and the European Leukodystrophy Association; and serves on the Board of Directors for the United Leukodystrophy Foundation and ALD Connect.

All other authors declare no relevant conflicts of interest.

## Abbreviations

ACMG/AMP –: American College of Medical Genetics and Genomics / Association for Molecular Pathology
aiALD –: Adrenal insufficiency in X-linked adrenoleukodystrophy
ALD –: X-linked adrenoleukodystrophy
AMN –: Adrenomyeloneuropathy
cALD –: Cerebral X-linked adrenoleukodystrophy
CHOP –: Children’s Hospital of Philadelphia
CI –: Confidence interval
ClinVar –: Clinical Variants database
FDA –: U.S. Food and Drug Administration
HR –: Hazard ratio
HSCT –: Hematopoietic stem cell transplantation
IQR –: Interquartile range
IRB –: Institutional Review Board
KKI –: Kennedy Krieger Institute
LC-MS –: Liquid chromatography–mass spectrometry
Loes score –: Magnetic resonance imaging severity score for ALD
LP –: Likely pathogenic
MGH –: Massachusetts General Hospital
MRI –: Magnetic resonance imaging
NBS –: Newborn screening
OR –: Odds ratio
P –: Pathogenic
P/LP –: Pathogenic or likely pathogenic
SU –: Stanford University
UM –: University of Minnesota
USA –: United States of America
UU –: University of Utah
VCEP –: Variant Curation Expert Panel
VLCFA –: Very-long-chain fatty acid
VUS –: Variant of uncertain significance

## Contributors Statement Page

Cecilie S. Videbæk and Keith P. van Haren conceptualized and designed the study and coordinated data collection. Cecilie S. Videbæk conducted parts of the statistical analyses, contributed to data collection, drafted the initial manuscript, and critically reviewed and revised the manuscript for important intellectual content. Keith P. van Haren supervised the study and provided overall guidance in study design and interpretation and critically reviewed and revised the manuscript for important intellectual content.

Danielle HJ Kim carried out the statistical analyses and critically reviewed and revised the manuscript for important intellectual content.

Zihuai He supervised the statistical analyses and critically reviewed and revised the manuscript for important intellectual content.

Hannah S. Hart, Robert Thompson, Razina Aziz-Bose, Lachelle Purnell-Savoy, Sonum Bharill, Ezzat Hashemi, Joseph Orsini, Elisa Seeger, Miranda McAuliffe, Isha Srivastava, Jennifer A. MacLean, Sejal Shah, Ali Fatemi, Julie S. Cohen, Eric Mallack, Troy Lund, Florian Eichler, Joshua L. Bonkowsky, and Laura Adang contributed to data collection, participated in interpretation of the data, and critically reviewed and revised the manuscript for important intellectual content.

Allan M. Lund supervised and provided guidance in interpretation and critically reviewed and revised the manuscript for important intellectual content.

All authors approved the final manuscript as submitted and agree to be accountable for all aspects of the work.

